# Projecting hospital resource utilization during a surge using parametric bootstrapping

**DOI:** 10.1101/2020.07.30.20164475

**Authors:** Jeffrey N. Chiang, Ulzee An, Misagh Kordi, Brandon Jew, Clifford Kravit, William J. Dunne, Ronald Perez, Neil R. Parikh, Drew Weil, Richard F. Azar, Robert Cherry, Karen A. Grimley, Samuel A. Skootsky, Christopher E. Saigal, Vladimir Manuel, Eleazar Eskin, Eran Halperin

**Affiliations:** Department of Computational Medicine, University of California, Los Angeles; Department of Computer Science, University of California, Los Angeles; UCLA Health, University of California, Los Angeles; Department of Radiation Oncology, University of California, Los Angeles; Department of Urology, University of California, Los Angeles; Clinical and Translational Science Institute, University of California, Los Angeles; Department of Family Medicine, University of California, Los Angeles; Department of Human Genetics, University of California, Los Angeles; Department of Anesthesiology, University of California, Los Angeles

## Abstract

During the initial wave of the COVID-19 pandemic in the United States, hospitals took drastic action to ensure sufficient capacity, including canceling or postponing elective procedures, expanding the number of available intensive care beds and ventilators, and creating regional overflow hospital capacity. However, in most locations the actual number of patients did not reach the projected surge leaving available, unused hospital capacity. As a result, patients may have delayed needed care and hospitals lost substantial revenue.

These initial recommendations were made based on observations and worst-case epidemiological projections, which generally assume a fixed proportion of COVID-19 patients will require hospitalization and advanced resources. This assumption has led to an overestimate of resource demand as clinical protocols improve and testing becomes more widely available throughout the course of the pandemic.

Here, we present a parametric bootstrap model for forecasting the resource demands of incoming patients in the near term, and apply it to the current pandemic. We validate our approach using observed cases at UCLA Health and simulate the effect of elective procedure cancellation against worst-case pandemic scenarios. Using our approach, we show that it is unnecessary to cancel elective procedures unless the actual capacity of COVID-19 patients approaches the hospital maximum capacity. Instead, we propose a strategy of balancing the resource demands of elective procedures against projected patients by revisiting the projections regularly to maintain operating efficiency. This strategy has been in place at UCLA Health since mid-April.

## Introduction

Preparations for a surge in COVID-19 patients in March and April in many hospitals included cancellation or postponement of elective procedures (Bedard et al. 2020), the expansion of the number of beds and ventilators in the hospital, and the creation of regional overflow hospital capacity. These preparations resulted in reduced hospital revenue and reduced access to care for non-COVID-19 patients. Specifically, many procedures that were delayed were medically necessary but not immediately urgent, resulting in a potentially negative effect on patient health. Fortunately, in almost all locations in the United States and in the world, the surge of COVID-19 patients did not overwhelm available hospital capacity, and hence in retrospect, if at the time there was a more accurate forecasting tool of future resource needs, the heavy price that was paid could have been avoided.

With the number of COVID-19 cases and hospitalizations increasing in many parts of the United States, healthcare systems are again making preparations for a possible surge. Yet, it is unclear whether a more optimal approach for the preparation of the health system may exist. Such an approach must rely on accurate predictions of resource capacity (e.g., hospital beds, ventilators) necessitated by COVID-19 patients in the near future. However, most models for hospital capacity utilized epidemiological models which were developed early in the pandemic. Because little data was available on actual hospital resource usage of COVID-19 patients, these models made assumptions that a constant fraction of patients would require hospitalization. (Weissman et al. 2020; Team, IHME COVID-19 health service utilization forecasting team, and Murray, n.d.; Moghadas et al. 2020; Shoukat et al. 2020; Wells et al. 2020). While these models were helpful for characterizing the pandemic at the population-level, they were not explicitly designed to aid in surge capacity planning at smaller scales (e.g., within municipalities or individual health systems).

In this paper, we demonstrate that the common assumption that regional case numbers are proportional to the hospital resources does not hold in practice, and therefore projections using this premise may be inaccurate. Furthermore, we propose a statistical method for prediction of necessary hospital resources for COVID-19 patients based on historical data found in the electronic health record. Our approach provides both predictions for the required resources in the near future, and confidence intervals for those predictions. Using a training and testing approach, we demonstrate that our approach provides accurate estimates and that the true level of resources needed falls within the confidence intervals. We also demonstrate that the approach is generalizable to environments that differ in the patient age, sex, or co-morbidity distributions.

Our approach, STOP COVID Hospital Projections, uses parametric bootstrap, where the model’s parameters are learned from the UCLA Health electronic health record (EHR). The model assumes a known trajectory of incoming patients, and predicts the effect of the trajectory on the resource allocation. Since we focus on short-term predictions (up to 3 weeks in the future), the range of possible scenarios in terms of the increase of the number of patients (doubling time) is limited. We thus make predictions under the worst-case scenario, as well as the most likely scenario (where the doubling time does not change). Using our approach, we show that there is no need to preemptively cancel elective procedures unless the actual capacity of COVID-19 patients gets close to the hospital maximum capacity. Instead, we propose a strategy of revisiting the projections every three days and unless the worst case projections show hospital usage close to capacity, continuing to schedule elective procedures as would be scheduled under normal circumstances.

Our method is implemented as a publicly available web-app at https://stopcovid19together.org/hospital. We hope this tool will be useful to healthcare systems as they plan how much capacity to make available for COVID-19 patients. In particular, since our worst-case predictions tend to only slightly overestimate the true needed resources (Figure 1), we believe that healthcare systems can use our worst-case predictions as a conservative approach to provide insight into the maximum amount of capacity that will need to be reserved for COVID-19 cases.

**Figure 1.**
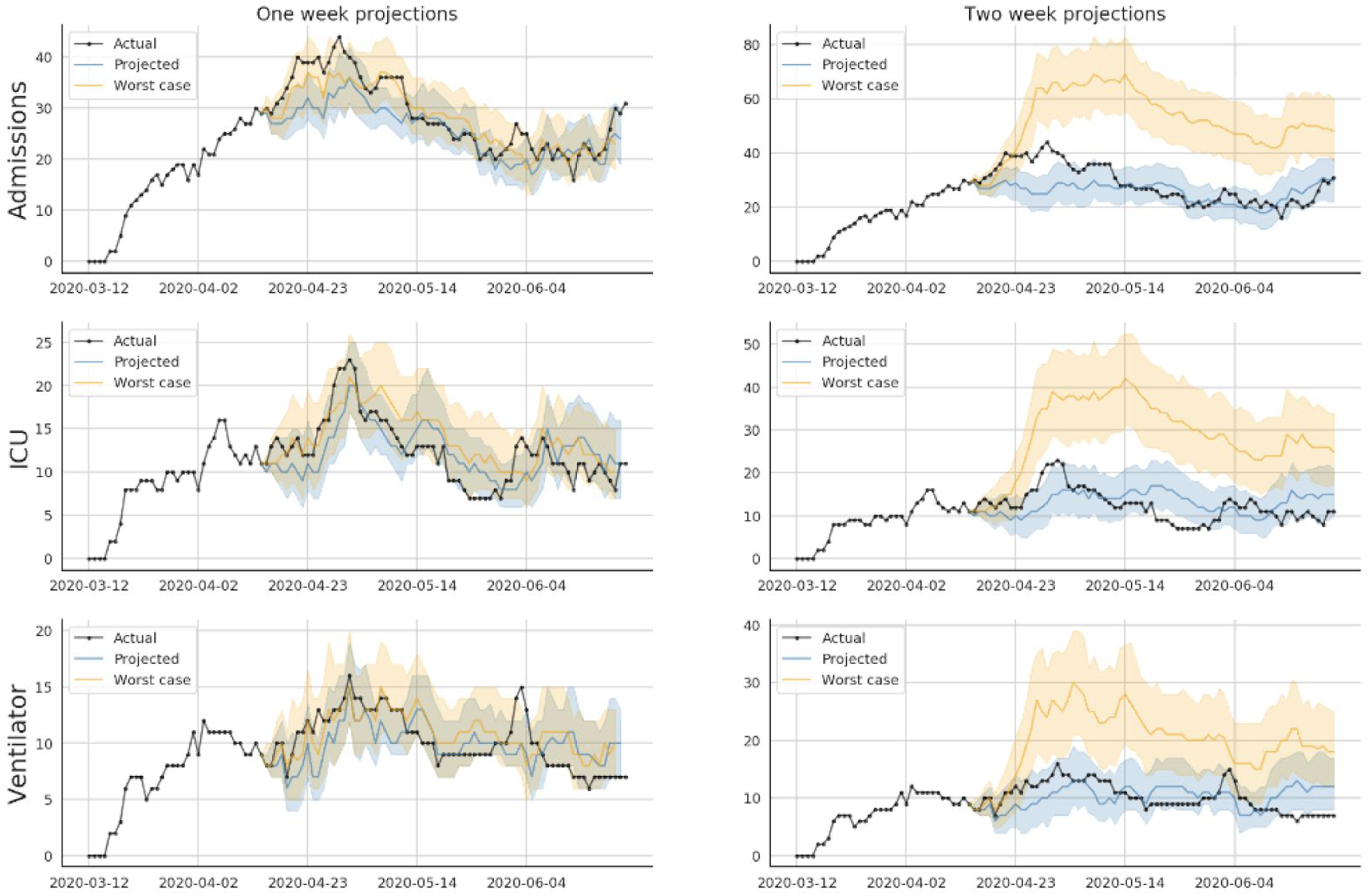
Comparison of model projections with observed cases at UCLA Health’s Hospitals. Validation of parametric model. The model was run on every day spanning 4/15/20 to 6/15/20. One and two week projections were recorded and aggregated. ICU and Ventilator cases are well captured by both one and two week projections. The spike of admissions in mid-April exceeded the model’s projections, however was still bounded by the worst-case scenario provided by the algorithm.

## Methods

### Parametric bootstrap model

Our approach is based on parametric bootstrap. Under this approach, we define a probabilistic generative model, which emulates the resource demand of COVID-19 patients. Input to our model are hospital census counts broken down by resource for the current day. For each individual in the hospital, we sample the patient’s specific resource usage from the distribution. We also sample the resource usage for each predicted new admission to the hospital. Each such sampling over all individuals provides a specific trajectory of resource demands over the duration of the prediction (e.g., two weeks). We repeat this process 1000 times, then take the median required capacity across all trajectories at each day as our prediction.

Below we describe the parameters and assumptions of this generative model. To generate hospital resource utilization trajectories, the model estimates the demand for resources by fitting a generative procedure over the reported features of patients to indicate resource use and duration of use. The distribution of patients (their parameters) is computed directly from the population and used as input when sampling trajectories from the model. Finally, the number of patients in future dates is assumed to follow exponential growth, where the exponent corresponds to the doubling time of the number of new cases. The doubling time can either be provided as an additional input to the algorithm, or otherwise it is estimated from the observed data (e.g., census counts) over the last two weeks. This approach allows for predictions under different growth scenarios, including a worst-case scenario where the doubling time is every four days.

### Patient Parameterization

The model operates over a distribution of patients, each characterized by a vector (*c*_1_,…, *c*_*n*_) of relevant covariates such as age, sex, and comorbidities. This vector can be directly sampled from observed data or from a distribution, which we assume to be multivariate Gaussian with mean vector *μ*_C_ and covariance matrix Σ_C_. These are estimated from the observed data by computing the sample mean and variance-covariance matrix, respectively.

### Resource requirement and duration

We assume that each individual *j* may potentially need resource-*r* (e.g., inpatient status, ICU status, or intubation). Thus, for each patient we assume the probability that the patient will use the resource *r* is given by 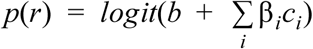. After determining the resource requirement for each patient, we assume that the amount of time spent using resource *r* (denoted by *l*_*j*_(r)) follows a Gaussian distribution truncated to be in a range of non-negative days:

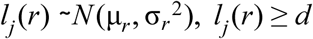

where *d* is the number of days the patient has already been using the resources. For new (simulated) patients, *d* = 0. We will assume that the expected duration μ_*r*_ is expressed as a function of the patient parameters such that 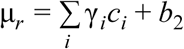. The set of parameters β_1_,…, β_*t*_, γ_1_,…, γ_*t*_, *b, b*_2_, σ_*r*_ are found using a maximum likelihood approach over the actual COVID-19 patients.

### Epidemiological Model

We assume that at day *t*, there are *n*_*t*_ new patients introduced to the population. Thevalue of *n*_*t*_ depends on the choice of epidemiological model, which is part of the input to our algorithm. The sequence *n*_*t*_ follows an exponential growth governed by doubling rate T_d_, i.e., *n* = *a*·2^*t*/ *Td*^. The parameter T_d_ can be estimated from previously observed case counts using a least-squares approach. That is, given a sequence of observed case counts (*n*_0_,…, *n*_*t*−1_), *n*_0_ > 0, the estimate for T_d_ is the value that minimizes the quantity 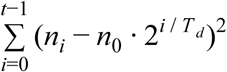. Alternatively, different values for T_d_ can be assumed and directly input to the model, particularly if one is interested to study the worst case scenario vs. the most likely scenario.

### Data

#### Observed data

Patient records were accessible through an extract of the UCLA Health electronic health records and retrospective data collection was approved by the University of California Institutional Review Board (IRB). For these analyses, relevant data for all patients with a positive COVID-19 polymerase chain reaction (PCR) test between March 1, 2020 and June 26, 2020, as identified by an internal registry, were used. For each patient, we extracted encounter-level information limited to their resource requirements (inpatient status, intensive care, intubation), as well as the duration of their usage (including admission date and total length of stay). We also extracted demographic information including age, sex, race, ethnicity, and smoking status for the same individuals. Encounter-level information was also extracted for elective procedures between January 1, 2019 and December 31, 2019 to summarize aggregate usage. Elective procedures were defined as scheduled surgical procedures (catalogued within an internal database) which did not originate from the emergency room.

#### Synthetic data

We used an oversampling procedure to construct two synthetic datasets, biased with a high mean age and low mean age respectively. In order to generate a dataset with artificially high mean age, we defined a patient pool as the observed patients. Patients over the mean age of the pool (46 years) were oversampled (replicated) 10 times to create a biased patient pool. Then, for each day between 6/1/20 and 6/22/20 (three weeks), we sampled from the biased pool the number of new cases that were actually observed that day. The same procedure was repeated for patients below the mean age of the pool to create a dataset with artificially low mean age.

## Results

### Hospital usage projections are well calibrated

We validated our approach with hospital resource usage data from UCLA Health. We evaluate the accuracy of our projections by applying our model to historical data. Specifically, we use data before a specific time point to predict the resource usage of the two weeks after the time point and compare our projections to actual hospital resource usage. Figure 1 shows our predictions compared to actual usage. As can be seen, the true number of patients using each of the resources are consistently within the predicted confidence intervals.

Our parametric bootstrap estimates the doubling time based on the previous two weeks. However, when there is a surge in cases, it is possible that the numbers of admitted patients will be increasing. We thus allow the doubling rate to be given as an input parameter to our model, allowing for a worst-case analysis as the surge approaches. Specifically, the historical worst sustained doubling time in the US was in New York in March, at a doubling time of every 4 days (Dong et al. 2020). We thus use 4 days as the doubling time, and we applied our model to generate worst case projections. In Figure 1, we demonstrate this approach when looking ahead one and two weeks. Using the parametric model, we see that the worst-case model captures the actual resource usage well within one week, but a 4 day doubling rate is clearly unrealistic when extrapolated to two weeks. However, it is important to note that the exponential functions predict similar near-term numbers.

### Projections can aid health system capacity planning

As shown in Figure 1, even the worst case projections in the near term only predict moderate growth in resource usage. Policies that preemptively increase capacity may drastically overestimate the number of COVID admissions which resulted in dramatic numbers of unused hospital resources. With a reliable model for predicting the increasing demands of COVID-19 patients, it becomes possible to balance the regular operating capacity against the expected surge of resource demands. Using historical data at UCLA Health, we used a bootstrapping procedure to simulate the effect of a sudden stop in elective procedures on operating capacity. We observed that resource requirements were generally reduced by half within two weeks, as shown in Figure 2. Thus, the projected resource usage due to COVID and the expected decline due to stopping elective procedures can be used to more efficiently balance a hospital’s operating load. Our simulations also show that the recovery of resources due to stopping elective procedures is faster than growth during moderate or even worst case surges (Figure 2).

**Figure 2.**
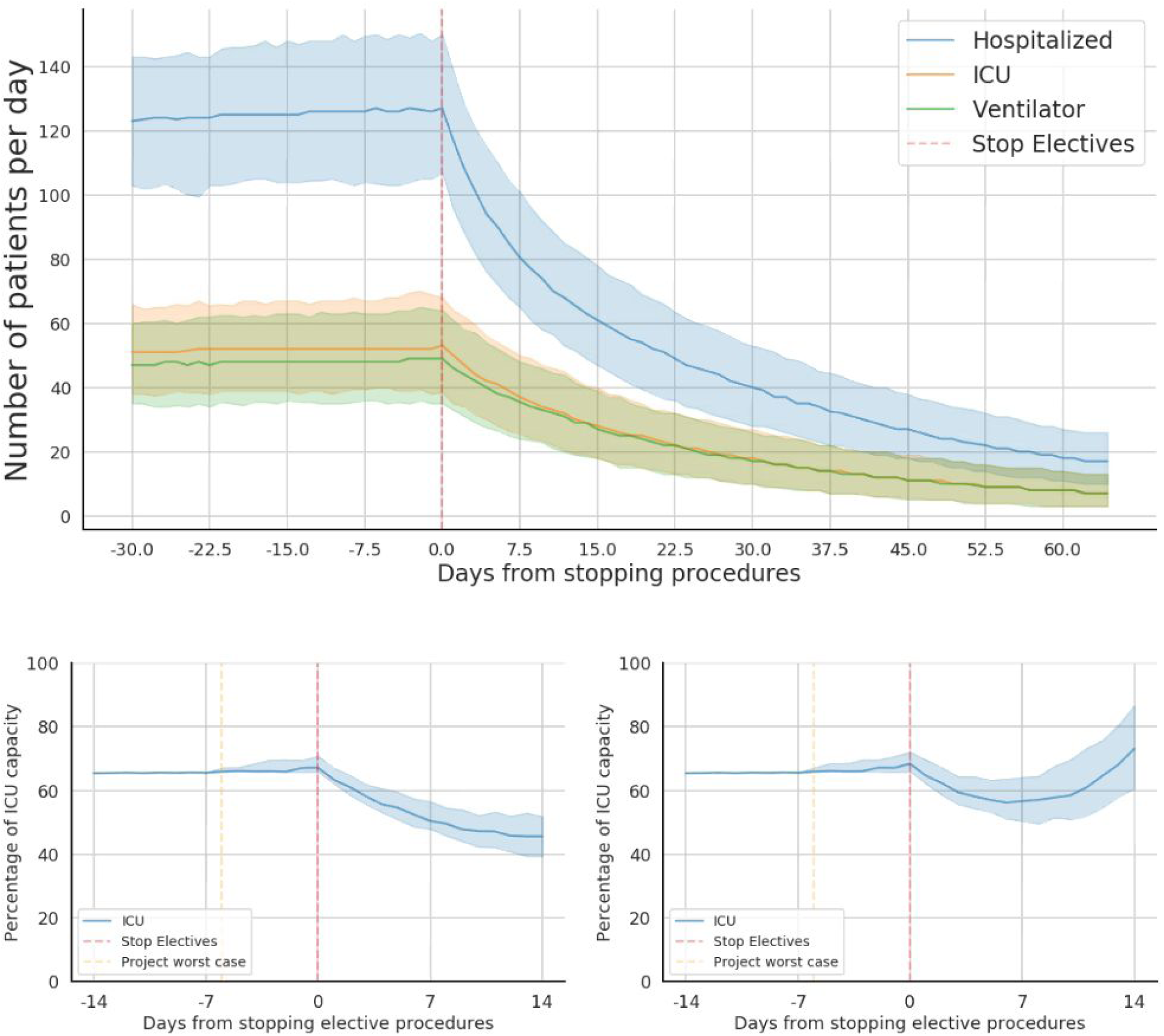
Balancing capacity against projected cases. Top. Decrease in resource demands due to cancelling elective procedures. This simulation begins assuming a standard operating load of 60 elective procedures per day. At the dashed red line, all subsequent elective procedures are canceled (i.e., there are no new elective procedures) and we observe the increase in operating capacity. Particularly, we find that the halving rate of the number of beds used by elective procedures is 15 days; that is, on average 50% of occupied resources can be freed up within 15 days. Bottom. Canceling elective procedures during a worst-case growth rate (doubling rate of [left] 8 and [right] 4 days). We assume that normal ICU capacity is around 70%. The stop in elective procedures is sufficient to accommodate a two week period with a sustained 4 day growth rate (right), and releases more resources than required in the case of a two week period with a sustained 8 day growth rate (left).

### Projections are robust for different patient demographics

One concern of any forecasting method is that each health system is different and has different demographics for patients. In this case, projections may be accurate for the health system that was used to estimate the parameters and inaccurate for other health systems. Specifically, the patient demographics may be different in different regions, resulting in different surge effects. We evaluated our projections in scenarios where populations with different demographic characteristics are admitted to the health system. To that end, we simulated data by a biased sampling of the UCLA Health population; we focused on a known risk factor for hospitalization, age, and tested the generalizability of our approach to a synthetic population with a bias towards higher and lower age. We generated populations with a mean ages of 65 and 30 by oversampling older and younger patients, respectively (see Methods). We then used the true COVID-19 patient data at UCLA Health before 6/1/20 to train our model and estimate its parameters. We applied the model on the simulated dataset, without accounting for the difference in population demographic characteristics. We observe that the parametric bootstrap in this case underestimates the required resources for the older population (Figure 3, left) and overestimates required resources for the younger population (Figure 3, right). However, if we provide the parametric bootstrap the mean age of the patients, the parametric bootstrap results in more accurate predictions of the required resources. We note that the mean value of each of the parameters of the patients (e.g. age, sex, comorbidities), is required as an input to our model; Figure 3 demonstrates that without this requirement, the estimates may be not be accurate. Finally, our illustration here focused on age, however our approach works in an equivalent manner on other demographic characteristics such as comorbidities, race and ethnicity, and sex.

**Figure 3.**
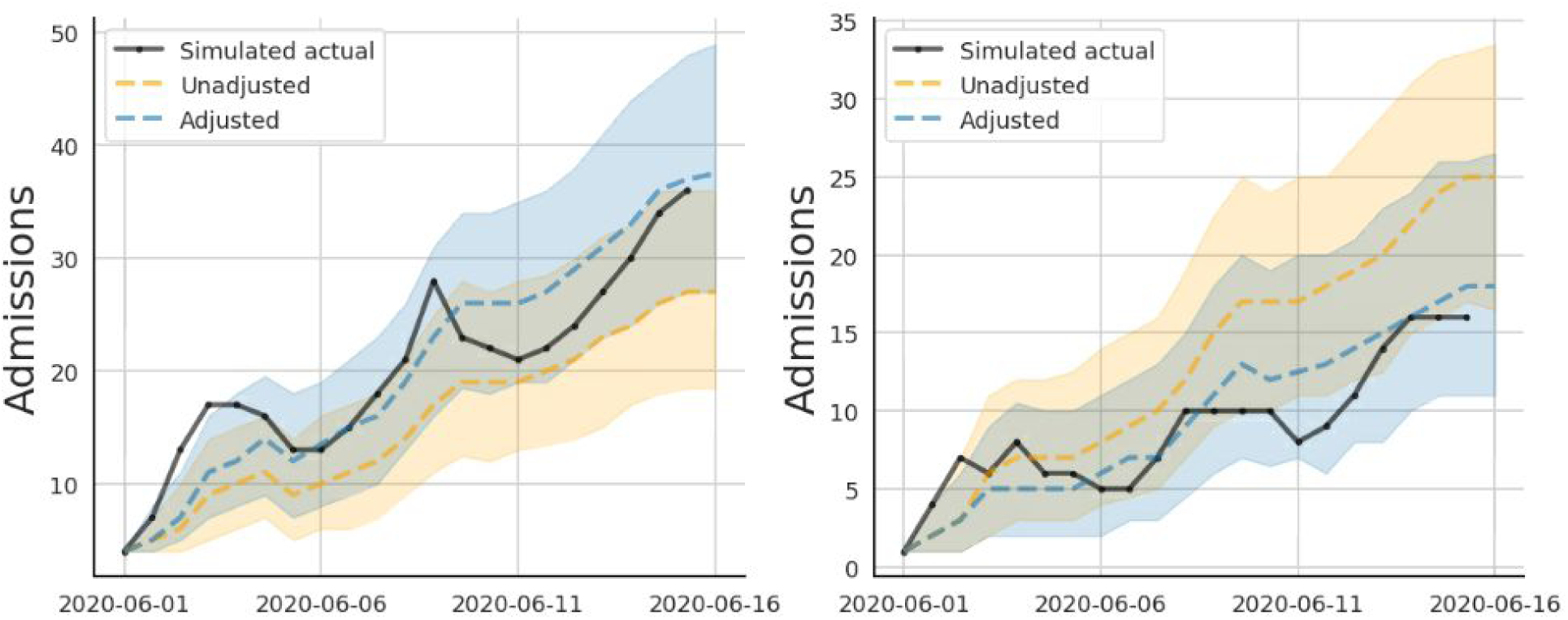
Simulations of different demographics. Left. Performance on a simulated population with higher average age. Right. Performance on a simulated population with lower average age. The dashed lines indicate the projected number of admissions and the solid lines indicate actual number of admissions. Orange: Projections without accounting for the difference in mean age. Blue: Accounting for the difference in age by providing the algorithm with the mean age of the patients in the health system. Our approach requires as an input the mean age of the patients in the health system, resulting in more accurate predictions.

### Near-term hospital usage projects are stable

In order to shed light on the factors that result in high accurate predictions, we hypothesized that most of the patients that are in the hospital are in the hospital for longer than a few days, and thus the near term predictions are dominated by the current patients. Indeed, by considering historical data in UCLA Health, we observe that the majority of the COVID-19 patients have been admitted for at least seven days (Figure 4).

**Figure 4.**
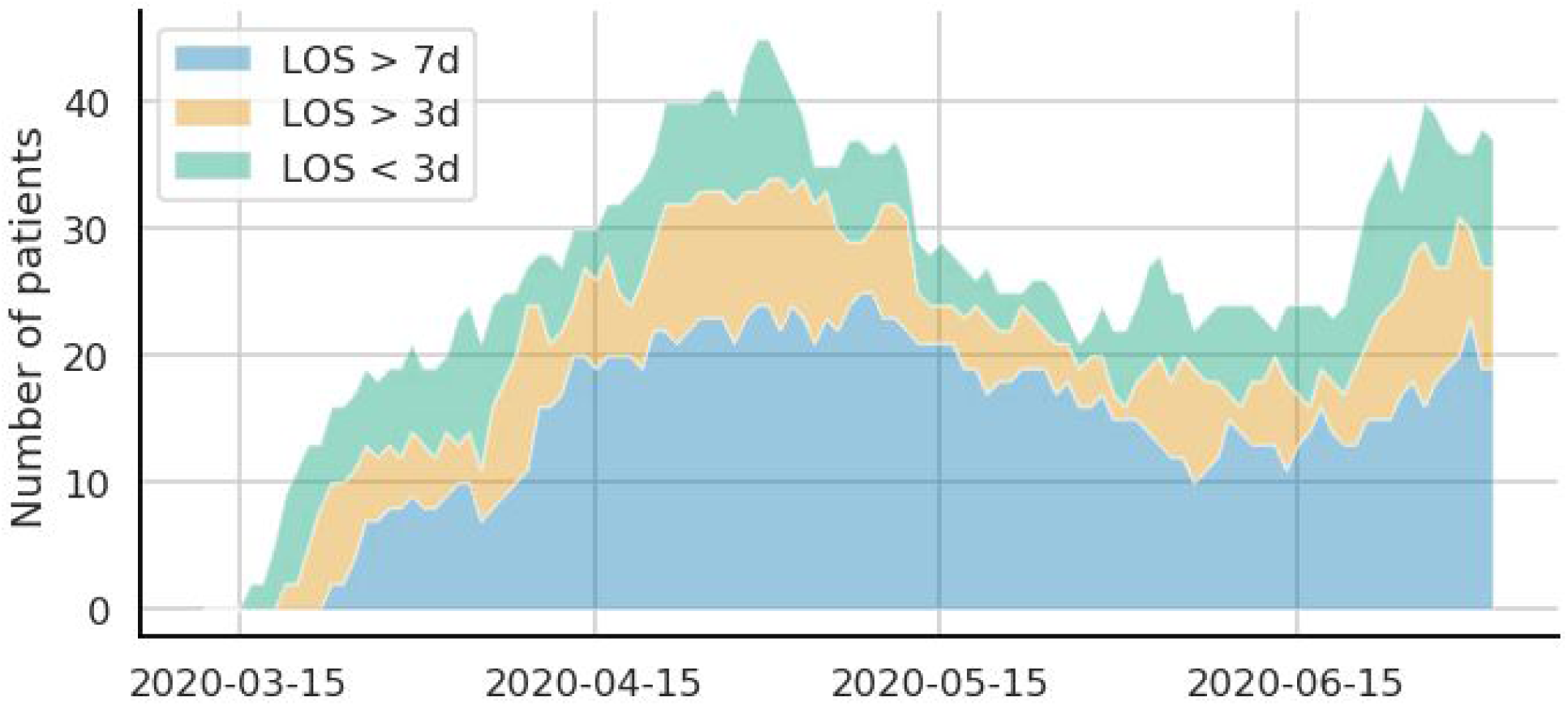
Length of stay composition over time. Blue: The number of patients who have been in the hospital for more than 7 days. Orange: Patients have been in the hospital for 3 to 7 days. Green: Patients who have been in the hospital for less than 3 days..

The projections themselves are a combination of two components. One component is the resource usage of COVID-19 patients already admitted and the other component is the resource usage or new patients. As can be seen in Figure 4, in the near term, the projections are dominated by the projected usage of current patients which can be accurately predicted.

### Diagnostic testing case counts are not a proxy for hospital usage

Current state-of-the-art approaches for hospital usage projections assume that a fixed fraction of incoming cases will require hospitalization and then use diagnostic testing data to estimate the current and future prevalence rates. However, diagnostic testing data is affected by ascertainment issues related to availability of testing, and policies around how tests are prioritized. The availability and policies have dramatically changed over time, and thus the numbers of positive diagnostic tests and the number of hospital admissions are only loosely related (Figure 5). Arguably, the success of our method stems partially from the fact that the approach presented here does not rely on this assumption, which is clearly violated in practice.

**Figure 5.**
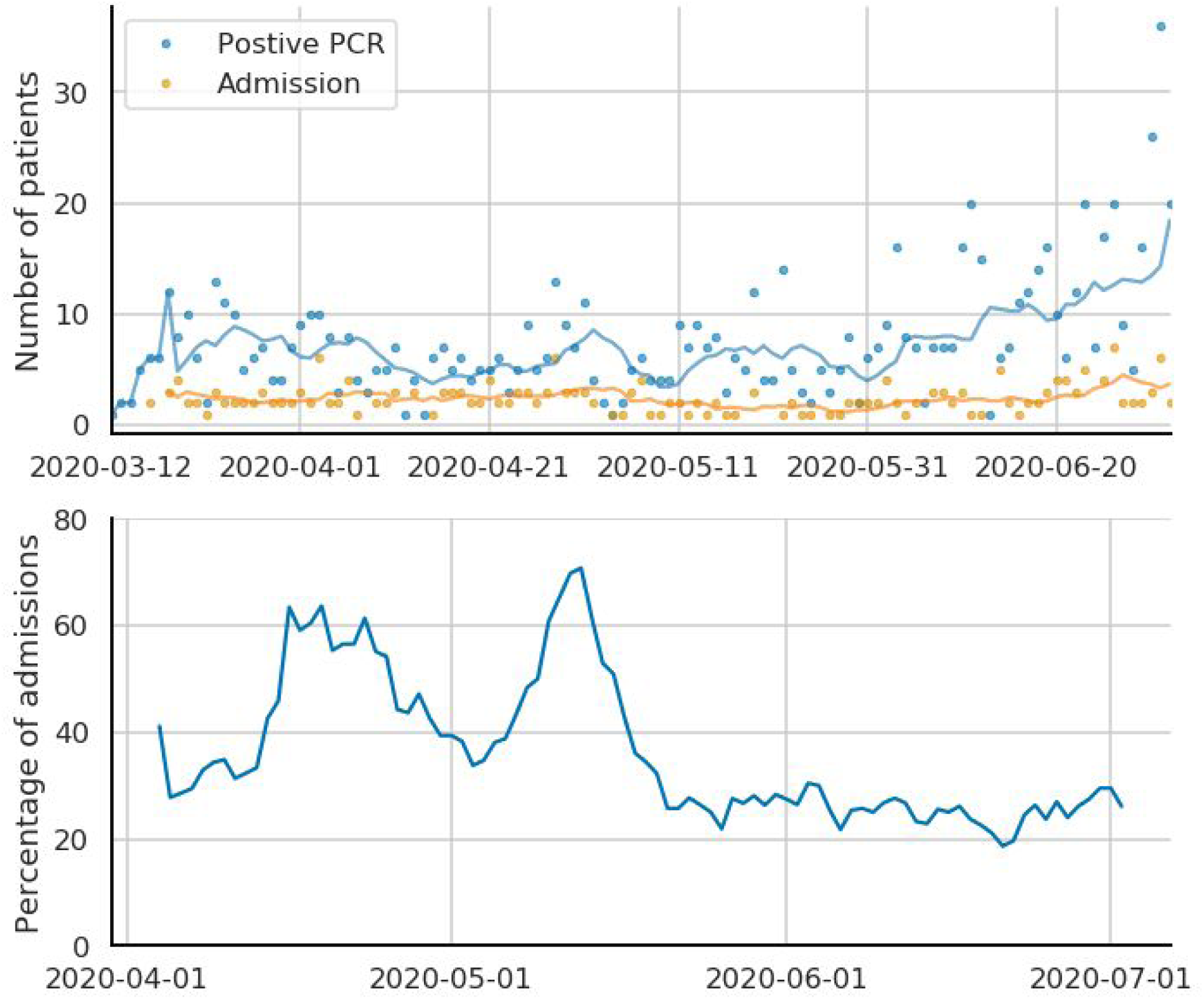
New positive COVID-19 cases and hospital admissions. Top. While new cases have increased recently, the number of new admissions has been relatively constant. Bottom. The proportion of patients testing positive that have converted (required hospitalization) has trended downward throughout the pandemic. This suggests that assuming a fixed proportion of cases will become hospitalized is suboptimal and will tend to overestimate cases as testing continues to become more available.

## Discussion

During the initial wave of the pandemic in the United States, public health authorities cautioned of a surge of high-resource COVID-19 patients, recommending a number of distancing and hygiene guidelines as well as opening hospital capacity. These recommendations were motivated by epidemiological projections as well as observations in locations where the hospital systems were overwhelmed. However, the assumption that hospital resource demand is directly linked to the epidemiological predictions is problematic (Figure 5). Greater availability of testing as well as clinical advancements have resulted in evolving care standards throughout the pandemic. For example, at the time of writing, clinical trials involving Remdesivir have been shown to slow the progression of disease (Beigel et al. 2020; Goldman et al. 2020) and there are new recommendations for the use of dexamethasone and other corticosteroids that show promise in reducing the time spent on supplemental oxygen (Yang et al. 2020; Horby and Nunn, n.d.). Similarly, better understanding of the transmission of the disease has led to changing clinical thresholds for intubation. In all of these cases, these treatments and decisions imply that the relative usage of limited resources (e.g. ventilators) will decrease throughout the course of the pandemic, implying that epidemiological models will continue to overestimate demand.(Beigel et al. 2020)

By leveraging our tool along with analysis of usage data from the UCLA Health system, we show several insights that can help guide surge planning for COVID-19 and future pandemics. First, for near-term projections (e.g., 3 or 7 day forecasts), even in worst-case surge scenarios, the majority of the projected hospital usage will be from patients that are currently hospitalized –enabling more accurate near-term forecasts. Second, since near-term forecasts are dominated by currently admitted patients, the hospital census indicators grow relatively slowly compared to other measures of the pandemic such as the number of positive case counts obtained from diagnostic testing. Third, using these accurate near-term forecasts, there is no need to preemptively cancel elective procedures unless the projected usage of COVID-19 patients approaches the hospital’s maximum capacity. Instead, we propose a strategy of regularly revisiting the projections, and adjusting the capacity available to elective procedures based on the projections. UCLA Health has been utilizing such a strategy since mid-April 2020. We note that in our experience, the alternative strategy of forecasting the surge months in advance has proven to be challenging, and resulted in less accurate forecasting as opposed to the model which prefers repeated, near-term adjustments.

In summary, we present a model for estimating resource demand that can be used to forecast the expected load on a hospital system during a pandemic-like situation. Our projection tool is also publicly available as a web-app at https://stopcovid19together.org/hospital so that other health systems can also use it for their projections.

## Data Availability

The data used in this study are not available for distribution due to patient privacy concerns.

